# Wastewater surveillance without prior lineage classification for reliable real time SARS-CoV-2 variant identification

**DOI:** 10.64898/2026.08.11.26359873

**Authors:** Raisa Kociurzynski, Sandra Reuter, Tjibbe Donker

## Abstract

The COVID-19 pandemic remains paradigmatic for the urgency of identifying emerging variants of rapidly mutating viruses in near real time. Grasping the infection dynamics enables better management of public health measures, including the timely allocation of resources. Wastewater surveillance has proven effective in estimating infection incidence and detecting variants in particular if testing rates declined due to milder disease manifestations. However, current methods typically rely on the prior classification of SARS-CoV-2 lineages or their signature mutations, which hampers the speed of variants detection. We present an alternative method that overcomes this limitation by identifying genetic changes in the viral population over time without requiring prior lineage classification. This approach was applied to wastewater samples from plants covering Swiss catchments in Altenrhein, St. Gallen, Geneva, and Zurich. To address noise, only samples with read depths above 40 and genome coverage of at least 90% were included. Genetic diversity within pooled populations over two time periods was compared to assess changes in viral composition. Application of this novel method enabled detection of shifts in genetic populations that corresponded to the emergence of known variants of concern and of the Omicron variant with reasonable precision without prior lineage classification. Notably the approach overcame the inherently high genomic noise in wastewater compared to clinical samples. In summary, we introduce a valuable tool for reliable, real time predictions for the emergence of potentially threatening virus variants from waste water samples that overcomes the need for prior lineage classification and high patient samples.

## 1 Introduction

The emergence of new viral variants has enabled SARS-CoV-2 to remain a cause of a considerable number of COVID-19 respiratory tract infections worldwide, despite the build-up of immunity against the disease in the population [3, 20, 27]. Given the importance of these variants to the epidemiology of COVID-19, robust yet affordable surveillance systems are warranted to track their emergence. As clinical testing and sequencing has decreased over time, surveillance systems based on submitted sequences from clinical isolates are increasingly at risk of no longer providing a reliable estimate of the course of the disease epidemiology [4, 24, 2]. As an alternative, wastewater surveillance has proven to provide a good indicator for incidence as well as the detection of variants of concern (VOCs).

However, wastewater samples tend to suffer from a low signal to noise ratio, creating specific challenges when using wastewater sequencing as a surveillance tool. For example, the library preparation of samples from wastewater poses difficulties as the RNA concentration might be low and PCR-inhibiting compounds might interfere with the amplification process. As a consequence, wastewater samples suffer from low sequencing coverage attributing to the low signal to noise ratio. Furthermore, the rate of the shedding of virus into wastewater for different variants might lead to a different abundance estimation as compared to estimations based on clinical isolates, even though this only plays into account when the transmission rates are decreasing or interventions are introduced [10].

A number of different methods aiming to amplify and disentangle the signal for predicting variants from wastewater have been introduced [25]. A widely used tool is Freyja, which recovers lineage abundance from mixed samples based on lineage deconvolution by solving a depth-weighted least absolute deviation regression problem [19]. For that, it uses a ‘barcode’ library with lineage-defining mutations generated from the resolved viral phylogeny based on clinical isolates.

Another approach employing variant-specific signature mutations could detect a local outbreak of Alpha almost two weeks prior to its detection in clinical samples as well as the first confirmed case of Omicron BA.1 [18][7]. Other methods relying on information from traditional clinical sampling include Kallisto, which is based on the principle used for transcriptome quantification to estimate lineage abundance, and uses multiple reference sequences per lineage to capture within-lineage variation [6].

However, such computational approaches relying on the a-priori classification of lineages cause the wastewater surveillance to still depend on clinical testing and sequencing information. This need for sequencing of clinical isolates is associated with considerable costs, threatening long-term implementation of the surveillance as well as impede effective use in lower- and middle-income countries. In addition, focusing only on lineage characterizing mutations can lead to cryptic mutations, which can be potentially important, remaining unnoticed [5].

Wastewater surveillance implementations that circumvent the use of clinical isolates may therefore have clear advantages if successfully able to timely detect variants. It has been demonstrated that sequencing of wastewater samples targeting the Spike receptor binding domain of SARS-CoV-2 or using a tiling amplicon-based approach can capture lineages that have not yet been identified from clinical sampling [13, 11]. Furthermore, it is possible to estimate variants based on changes in the genetic population diversity over time without prior lineage classification [9], by detecting shifts in the genetic population structure estimated from collected clinical isolates. Theoretically, such a method should also be applicable to wastewater genomic data alone making wastewater surveillance far more efficient and effective in detecting emerging viral variants by eliminating the need for clinical isolates. Here, we thus adopt this population genetic diversity-based method to wastewater genomic data, and apply it to openly available sequencing data obtained from three wastewater plants in Switzerland.

**Table 1:**
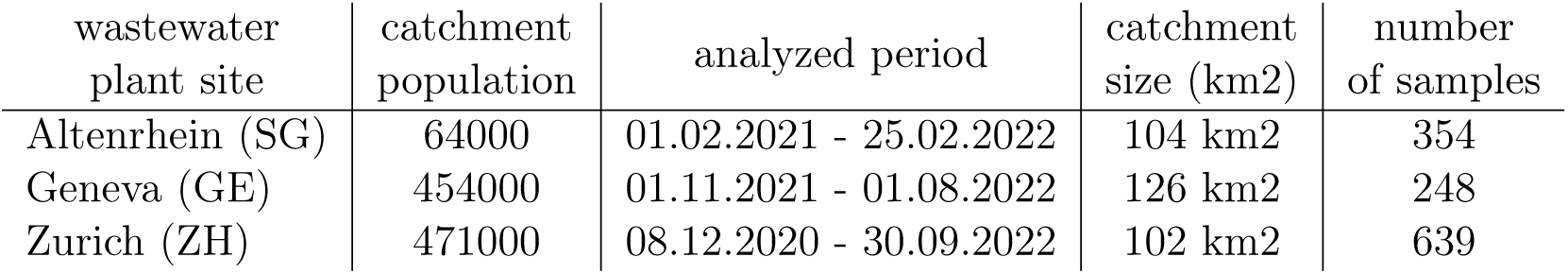
Wastewater plants from which samples taken were analyzed. The assigned catchment population, analyzed time period, catchment size, and sample size are given. The wastewater facility in Altenrhein is assigned to a catchment in St. Gallen (SG).

## 2 Methods

### 2.1 Data

Sequenced samples from wastewater treatment plants from three Swiss sites located in Geneva, Altenrhein (St. Gallen), and Zurich taken from the ENA Project PRJEB44932 were analyzed. For the analyzed time periods a total of 248 samples for Geneva, 354 for Altenrhein and 639 for Zürich were available. Only complete and high coverage samples, with entries below 1% Ns and *<* 0.05% unique amino acid mutations, were selected. The abundances of lineages from wastewater were estimated by generating a deconvolution matrix as implemented within the Freyja method taking the trimmed cram files filtered by coverage with a cut-off of 70 as input [19]. Table 1 summarizes the corresponding population catchment sizes of the wastewater facilities and timelines for the deployed samples. During these time periods samples are available at a near daily basis.

Incidence data based on SARS-CoV-2 RNA concentration in wastewater and on new clinical COVID-19 cases in the three catchment areas was retrieved from the website of the Swiss Federal Institute of Aquatic Science and Technology (Eawag) [1]. Data for the distribution of SARS-CoV-2 lineages from clinical samples over time was either downloaded from the Global Initiative on Sharing All Influenza Data (GISAID) [22] directly or through the Outbreak.info R package [26, 12].

### 2.2 Bioinformatics pipeline

Sequences were downloaded from the European Nucleotide Archive (ENA) in an aligned cram format. In cases where samples were sequenced using the targeted amplicon approach via the ARTIC protocol [21], primers were trimmed using the iVar trim tool [14] depending on the primer scheme stated in the metadata. Variant calling was performed with the HaplotypeCaller tool from the GATK package [8] using the original SARS-CoV-2 reference genome (RefSeq ID NC_045512) from NCBI [28]. Bases with a Phredscaled quality score below 10 were discarded. Genome coverage and mean sequencing read depth were calculated using the samtools coverage tool. Further analysis was performed either on a unfiltered set of samples or only filtered samples with a read depth higher than 30 and a coverage of more than 90%.

### 2.3 Variant detection

Figure 1 illustrates the implemented pipeline used to estimate emerging variants and their abundance. As a measure for the genetic population diversity between two different time points we deployed the logK value as previously introduced [9]. An increase in the genetic population diversity, and therefore logK, is indicative for the take over of a new variant with a higher fitness advantage compared to the previously predominant lineage. To show the progression of logK over time we correlated logK with time over time windows of the previous 21 and 42 days using the ranked Kendall’s Tau correlation. Thereby the correlation was performed for logK values calculated between a reference date and each date in the time window. The p-values for those correlations are plotted over time. We define the emergence of a new variant when the *p*-value is below a threshold of 0.05 divided by the number of data points as it is applied in the Bonferroni correction. For comparing the first day of detection to clinical patient data we first define periods of predominant variants. Thereby, the first day of detection of a variant in clinical patient data is when its abundance reaches 5% whereas the predominant period of this variant ends when its abundance reaches 95%. In this way we define the first detection of a variant in wastewater when its smoothed p-values first cross our defined threshold and this date lies within the clinically defined period for the predominant variant. Additionally to detections based on smoothed p-values we note single point estimates that cross the threshold within the defined period for the predominant variant.

**Figure 1:**
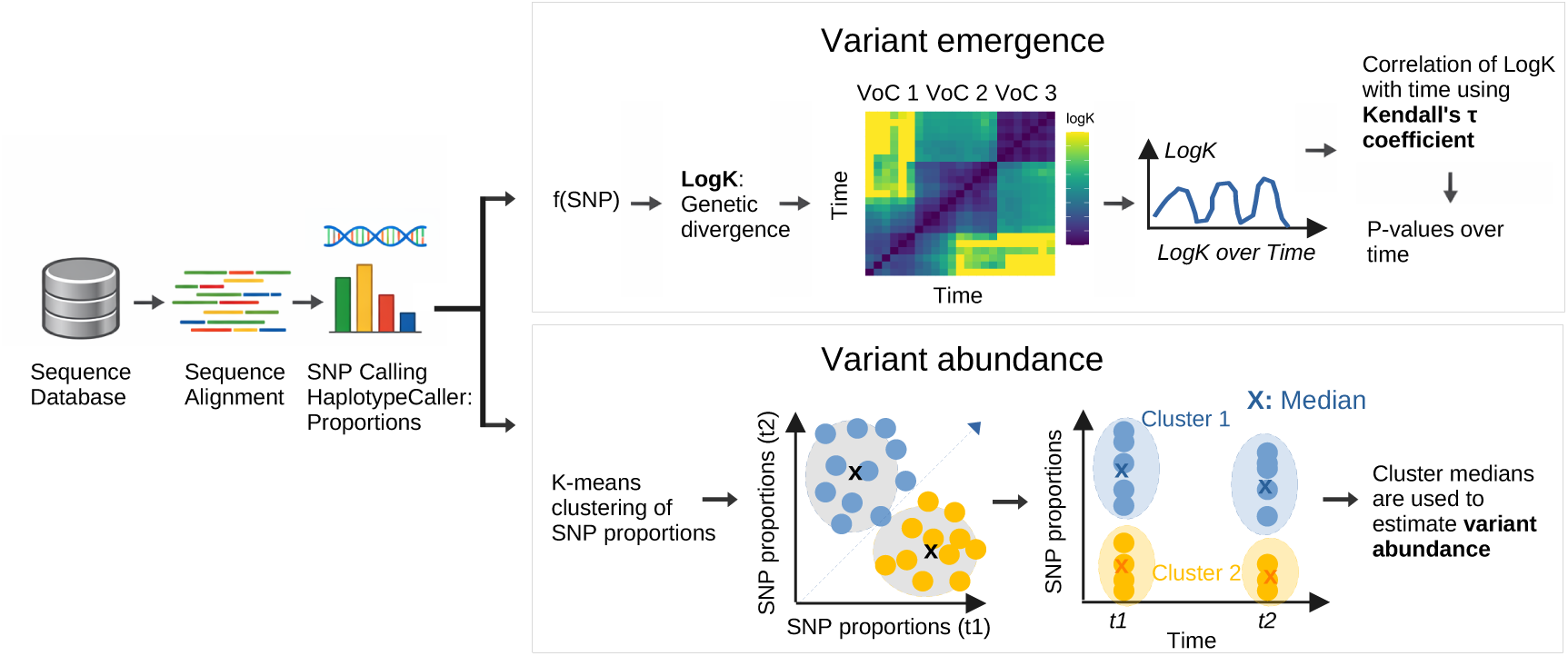
Schematic representation of the algorithm of variant estimation. COVID-19 sequences from wastewater samples are aligned to a reference sequence and the proportions of SNPs are estimated. logK is calculated pairwise based on the SNP proportions in two samples, illustrated as a heatmap. logK over time can be plotted as the median logK difference over a sliding time window.

### 2.4 Abundance estimate

The abundance for the variants was calculated by clustering all SNPs with the k-means clustering method using *k* = 2. To investigate if data quality influences our abundance estimate, a ranked Kendall’s Tau correlation was calculated between residuals of our estimate and the quality control parameters read counts, coverage, and read depth. The residuals were defined as the absolute difference between the moving average of the estimate over time windows of 25 days and our estimate at the corresponding time point.

## 3 Results

### 3.1 Variants as changes in logK

A pairwise comparison of the SARS-CoV-2 genetic populations structure of all included wastewater samples shows clear groups of similar populations visible as blocks of low logK values surrounded by high logK values in the heat maps (Figure 2). Although variability within the groups exist, transition to higher logK values happens relatively quickly, resulting in a clear separation between the groups, indicating temporal separation between distinct variants. The observed transitions generally correspond to dates of transition between known VOCs.

**Figure 2:**
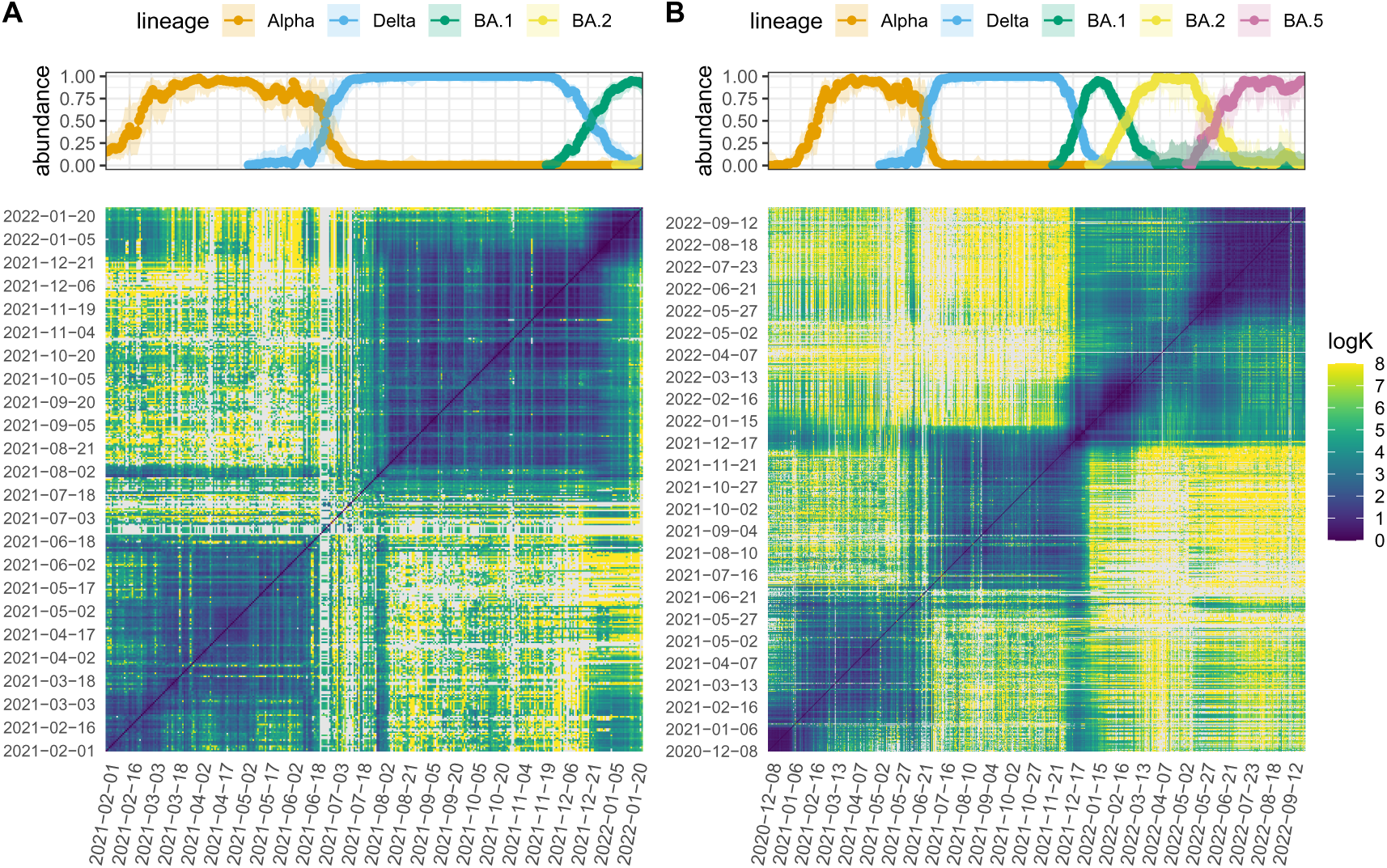
logK estimates on a nearly daily basis from wastewater in (A) Altenrhein, St Gall and (B) Zurich for all date combinations. Above the abundance of VOCs based on clinical case data is shown. Abundance estimates are shown only for days on which wastewater samples are available and displayed on a discrete time scale. Clinical case data was obtained from GISAID.

In Altenrhein, we detect changes in the genetic structure corresponding to dates of the transitions from the Alpha to Delta variant and from the Delta to the BA.1 variant (Fig. 2A), while for Zurich three distinct blocks with similar genetic footprints emerge at the onsets of Alpha, Delta, and Omicron, respectively (Fig. 2B). Here, BA.1 and BA.5 are clearly distinguishable while the BA.2 signal is less obvious, likely because the Omicron sublineages evidently share many SNPs (Fig. 2B). A similar pattern is observed in Geneva, where the transition to BA.2 was not captured, which here may also be influenced by insufficient availability of wastewater samples during this time period (Fig. SS4).

### 3.2 Correlation of logK with time

To be able to track transitions over time we performed ranked Kendall’s Tau correlations between logK and time. The p-values of these correlations are employed as a measure for the progression of the variant, where small p-values represent strong correlations indicative of an increase or decrease in logK pointing to an emergence of a new variant. The -log(p-value) for the correlation between logK and time peaks at distinct times, generally overlapping with the emergences of new variants. We observe two peaks in March and April 2021 for Altenrhein, which fall into the time period of the final establishment of the Alpha variant (Fig. 3A), with the first peak starting on the 11th of March 2021 and corresponding to an abundance of already 74% of Alpha in clinical samples. In Zürich, only a small, non-significant peak in p-values is observed during the emergence of Alpha, likely because the number of data points in the beginning of the time series is not sufficient to establish a significant correlation (Fig. 3B).

**Figure 3:**
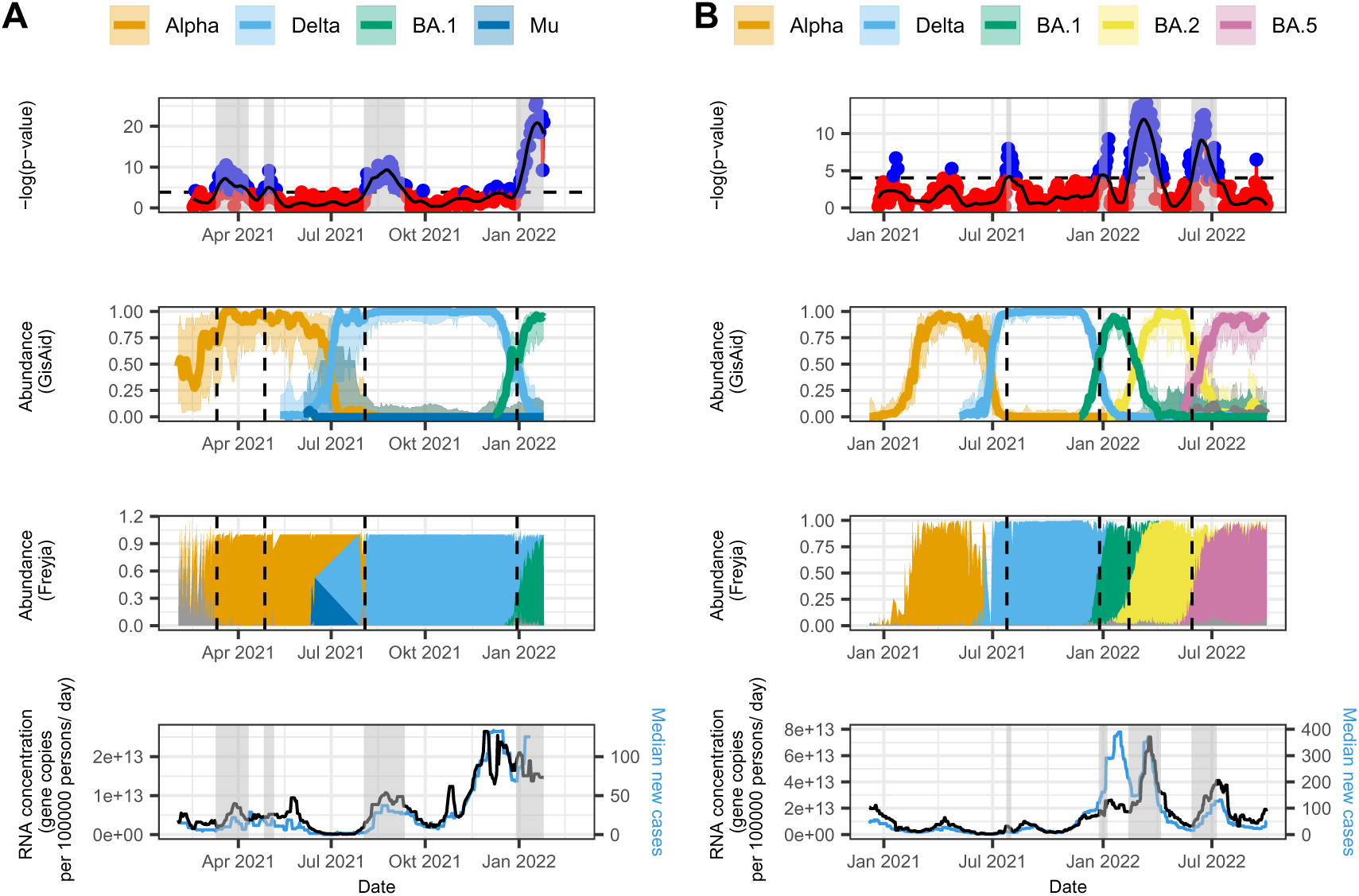
Variant and incidence estimates for (A) Altenrhein, St. Gallen and (B) Zurich. Upper panel: P-values for logK ranked correlations with time for periods of 42 days. The black line represents loess smoothing. Blue dots and grey transparent areas represent values that are above the detection threshold (see Methods section, dashed horizontal line). Second panel: Abundance of VOCs based on clinical samples obtained from GISAID. Third panel: Estimation of VOCs in local wastewater using Freyja. Lowest panel: Sars-CoV-2 RNA concentration in the local wastewater facility as a function of time and the median local incidence.

The emergence of Delta is observed in Altenrhein starting on the 3rd of August 2021, and in Zürich on the 25th of July 2021. While the transition to Delta in clinical samples is detectable as early as June 2021 in both places, our analysis fails to detect Delta at its growing phase possibly due to low number of reads and low coverage of the sequences during that time period (Fig. SS1 and SS3).

The emergence of Omicron BA.1 coincides with the strongest signal in our analysis. The strongest signal in the Altenrhein catchment is observed at 30th of December 2021, which coincides with the arrival of the Omicron BA.1 variant. On this date BA.1 had a proportion of approx. 54% in the St. Gallen area based on clinical isolates deposited in GISAID (Fig. 3A). In Zurich the signals for BA.1 (starting at the 26th of December), BA.2 (starting at the 13th of February), and BA.5 (starting at the 29th of May) are significant and clearly distinguishable, consistent with their emergence patterns observed in clinical case data (Fig. 3B).

We furthermore observe a significant change in the SARS-CoV-2 genetic population structure within the Geneva catchment starting from the 2nd of December 2021 (Fig. SS5A, Table 3). Interestingly, this is 2 days prior to when the first BA.1 sublineage was sequenced in a clinical sample in Geneva. Strikingly, by analyzing the lineage defining barcodes in the beginning of this peak we determined the Delta sublineage AY.43 and not BA.1 as the predominant lineage. A second peak crossing the detection thresh-old at the 15th of February 2022 overlaps with the emergence of BA.2 in clinical cases, with about 17% of sequenced samples being assigned to BA.2 at this point in time. The Geneva analysis further reveals a peak that corresponds to the emergence of BA.5 exceeding the threshold at 10th of May 2022, a time when BA.5 only accounted for around 2% of all clinical samples (Fig. SS5)

### 3.3 Comparison with Freyja

We compared our results with VOCs predicted by Freyja, which is considered to be the gold standard method for variant estimation from wastewater. In Altenrhein, Freyja detects Alpha from the start of our measurement at the 1st of February 2021 and therefore over a month prior to us. Notably, the period before the 11th of March shows some noise with other additional lineages estimated. Our method estimates the introduction of Delta into the St. Gall population in late July, slightly later than indicated by both clinical testing and estimates obtained with Freyja (Fig. 3B, Table 3). The low data quality during this time might also explain why Freyja estimated the lineage Mu on three days in June with an abundance reaching 52% while Mu does not exceed an abundance of 6% in clinical sampling. As mentioned above, BA.1 in Altenrhein was first detected at the 30th of December. The estimated abundance for BA.1 from Freyja on this date was approx. 19% based on the corresponding wastewater samples (Fig. 3A).

The emergence of BA.1 in Zurich is estimated to take place at 26th of December 2021, 18 days later than predicted by Freyja. At this point in time BA.1 has an abundance of 59% based on sequenced cases. We estimate the transition between BA.1 and BA.2 in Zurich to happen at the 13th of February 2022, which corresponds to an abundance of 27% for BA.2 based on clinical sampling, very close to the abundance estimated by Freyja (28%). The emergence of BA.5 is estimated at the 29th of May 2022, 20 days later than the prediction made by Freyja. The clinical abundance of BA.5 on its estimated onset was 24%.

### 3.4 Sensitivity analysis

#### 3.4.1 Effect of reduction of sampling frequency

When the sampling is performed daily Alpha is first detected on the 11th March 2021 in Altenrhein which falls within the period when Alpha is clinically predominant. The first day of the first detectable peak that is closest to the emergence of Delta in patient samples is the 3rd of August 2021 when sampling daily which is 25 days after the end of the clinical Delta period (Fig. 5A and Table 2). Reducing the sampling frequency to three times a week in Altenrhein results in the failure to detect Alpha even though single point estimates cross the detection threshold within the predefined time period (Fig. 5A and Table 2). Furthermore, sampling three times a week leads to a detection of Delta and BA.1 six days later than with daily sampling. Similarly as for daily sampling, Delta is detected outside the time period where it was the clinically predominant variant in Altenrhein. When sampling on a weekly basis BA.1 is the only variant that is detected in Altenrhein.

**Figure 4:**
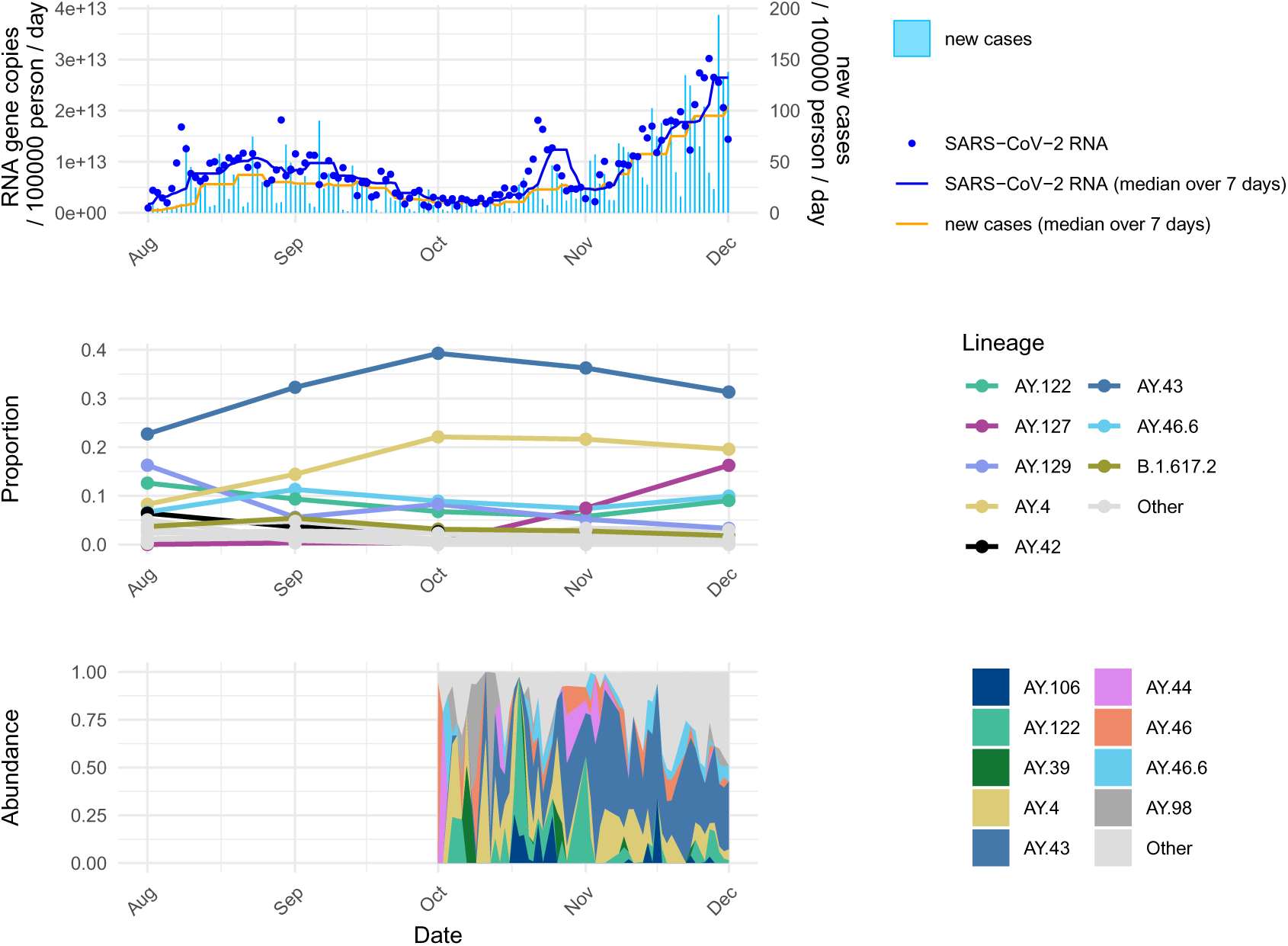
Analysis of the October incidence peak in Altenrhein observed based on wastewater samples. In (A) the incidence is represented based on RNA gene copies of SARS-CoV-2 per 100000 people per day in wastewater. On the right y-axis new cases per 100000 people per day in St. Gallen are shown. In (C) the lineage proportions for clinical samples for Delta and its sublineages for the whole of St. Gallen is depicted (data obtained from GISAID). In (B) the lineage distribution for samples from the Altenrhein wastewater plant between October and December 2021 is shown. Variant abundance was estimated using Freyja.

**Figure 5:**
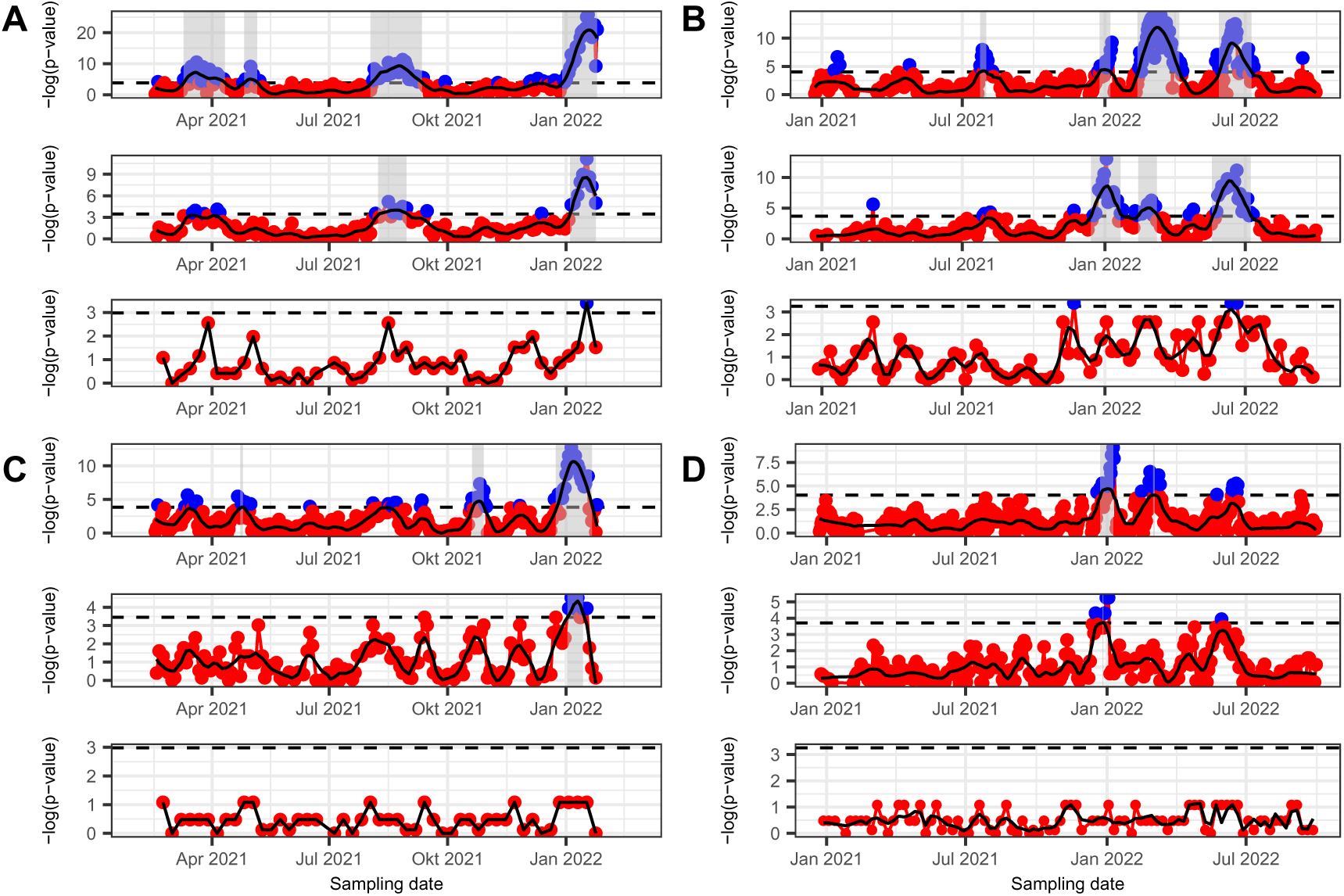
Sensitivity analysis of p-values for the logK correlations for (A and C) Altenrhein and (B and D) Zurich. The correlations were performed over 42 (A and B) and 21 (C and D) days. The panels for each subplot represent correlations for samples taken daily (upper panel), three times a week (middle panel), and weekly (lower panel). The dashed horizontal line represents the threshold for detection (see Methods section).

**Table 2:** First detections of VOCs in Altenrhein wastewater. Columns “first date” and “last date” represent the start and end day of the predominant variant or lineage defined based on clinical data (See method section for details). The remaining columns show the days of detection when sampling is carried out daily, three times a week, or weekly. Estimated detection dates are based on p-values for logK correlations of 42 days

| lineage | first_date | last_date | daily_offset42 | MoWedFri_offset42 | weekly_offset42 | daily_point_esti | MoWedFri_point_esti | weekly_point_esti |
| --- | --- | --- | --- | --- | --- | --- | --- | --- |
| Alpha | 2021-02-16 | 2021-03-17 | 2021-03-11 | NA | NA | 2021-02-18 | 2021-03-17 | NA |
| Alpha sublineage (?) | NA | NA | 2021-04-27 | NA | NA | NA | NA | NA |
| Delta | 2021-06-10 | 2021-07-09 | 2021-08-03 | 2021-08-09 | NA | NA | NA | NA |
| Omicron: BA.1 | 2021-12-11 | 2022-01-18 | 2021-12-30 | 2022-01-05 | 2022-01-17 | 2021-12-11 | 2021-12-13 | 2022-01-17 |

Similar as in Altenrhein, the detection of Delta in Zurich is delayed by 11 days regarding the defined period for Delta. The Omicron lineages BA.1, BA.2, and BA5 are all detected during the time periods when these VOCs are clinically predominant (Fig. 5B and Table 3). In Zurich, Delta is only detected when testing is performed daily. Notably, BA.1 is detected 11 days earlier when testing is performed three times a week than it is captured when using a daily testing strategy (Fig. 5B and Table 3).

**Table 3:** First detections of VOCs in Zurich wastewater. Columns “first date” and “last date” represent the start and end day of the predominant variant or lineage defined based on clinical data (See method section for details). The remaining columns show the days of detection when sampling is carried out daily, three times a week, or weekly. Estimated detection dates are based on p-values for logK correlations of 42 days

| lineage | first_date | last_date | daily_offset42 | MoWedFri_offset42 | weekly_offset42 | daily_point_esti | MoWedFri_point_esti | weekly_point_esti |
| --- | --- | --- | --- | --- | --- | --- | --- | --- |
| Alpha | 2021-01-23 | 2021-03-30 | NA | NA | NA | 2021-01-24 | 2021-03-08 | NA |
| Delta | 2021-06-08 | 2021-07-14 | 2021-07-25 | NA | NA | NA | NA | NA |
| Omicron: BA.1 | 2021-12-02 | 2022-01-24 | 2021-12-26 | 2021-12-15 | NA | 2021-12-23 | 2021-12-17 | NA |
| BA.2 | 2022-01-24 | 2022-04-02 | 2022-02-13 | 2022-02-14 | NA | 2022-02-14 | 2022-01-24 | NA |
| BA.5 | 2022-05-17 | 2022-07-19 | 2022-05-29 | 2022-05-20 | NA | 2022-05-24 | 2022-05-25 | NA |

Interestingly, in Geneva BA.1 is estimated 11 days earlier in wastewater than in clinical samples. As the BA.1 signal does not fall below the detection threshold, we identify the start of the subsequent peak at the trough separating it from the preceding peak. The onset of this peak occurs within the clinically relevant BA.2 period. Similar as to BA.1, we estimate BA.5 six days prior to when it is observed in clinical data. In Geneva, reducing the sampling frequency to three times a week still leads the detection of BA.1 five days earlier than the day it first crosses the detection threshold in clinical isolates (Fig. SS7A and Table S3). Concurrently, sampling wastewater three times a week leads to an estimate of BA.2 that lies within the defined time period for Geneva. BA.5 is estimated on the exact same day as the onset in patient data. Weekly sampling fails to estimate any variant. However, despite the strong noise it is possible to estimate BA.1 and BA.2 in the weekly data subset based on single estimated data points.

#### 3.4.2 Effect of correlation time

Overall, reducing the correlation time from 42 to 21 days leads to less defined signals and more false positive peaks. In Altenrhein, the shorter correlation time leads to a delay in the detection of Alpha and Delta of 44 and 79 days, respectively, when using daily sampling (Fig. 5C and Table 1). As we observe no signal within the clinically relevant time ranges for the variants, the detection was estimated from the peaks crossing the threshold that are nearest to the variant’s specific periods. Interestingly, single point estimates can be observed during the periods of clinically predominant Alpha and Delta (Table 1). Nonetheless, BA.1 can be detected with both sampling strategies within the expected time period.

In Zurich, the reduction of the correlation time to 21 days results in only BA.1 and BA.2 being detected when sampling daily while only BA.1 generates a detectable signal when taking samples three times a week (Fig. 5D and Table 2). The times of these detections lie within the predefined time ranges for the VOCs based on clinical samples. In Geneva additional peaks emerge when a 21-day correlation period is employed (Fig. SS7B). Strikingly, by analyzing the lineage defining barcodes in the beginning of this peak we determined the Delta sublineage AY.43 as the predominant lineage, the same lineage that was observed during the October peak in Altenrhein.

By examining the first two peaks it becomes apparent that the large peak which is observed from the 42-days correlation analysis in fact constitutes two peaks combined into one, namely the AY.43 Delta sublineage peak and the BA.1 peak. This sublineage is represented as a small peak. While several peaks most likely stem from Omicron sublineages when sampling daily, a peak with an onset within the period of clinically predominant BA.2 can be detected. We also monitor a peak that is representative of BA.5 and reaches the detection threshold 6 days before the BA.5 abundance in clinical samples passed 5% (Table S4), similarly as we see for the 42-day correlation period. When the data is subset to reflect three times a week sampling, BA.1 and BA.5 are detected, whereas BA.2 is not.

### 3.5 Sublineage detections

As reported above, using the shorter correlation time window we monitor an additional peak in the St. Gallen area that crosses the detection threshold but does not represent the main circulating VOCs at that time (Fig. 5C). Data on incidence based on SARS-CoV-2 RNA concentration from the wastewater plant in Altenrhein, representing a part of the catchment area of St. Gallen, shows a similar peak spanning a time period from 2021-10-18 to 2021-11-01, with the highest value observed on October 22nd (Fig. 4A). Interestingly, no peak is observed in the incidence of new Covid cases based on clinical samples for the same catchment area (Fig. 4A). By further examination of the lineage distribution from clinical sampling we identified the two Delta sublineages AY.4 and AY.43 (together with its sublineage AY.43.4) to be the predominant lineages in the complete St. Gallen area (population of 76328) in October 2021 (Fig. 4C). Notably, the sublineage AY.43 also represents the predominantly circulating lineage that, in our analysis, yielded a signal in Geneva in early December. For comparison, we analyzed the circulating Delta sublineages in the Altenrhein wastewater plant using the Freyja lineage deconvolution method on daily sampled probes from the 8th to the 28th of October 2021. Lineages AY.4 and AY.43 combined with their sublineages again showed a high prevalence (Fig. 4B). When assigning SNPs to barcode defining lineages and ranking them, we observe AY.43 to be the main circulating lineage over most of October and November, in line with preceding results.

### 3.6 Incidence

A juxtaposition of the p-values with the incidence based on wastewater RNA concentration and on new cases (Fig. 3) is in good overall agreement, with our logK analysis covering the most prominent peaks seen in the incidence. In Altenrhein, a noticeable peak at the end of October 2021 that exists in the RNA concentration in local wastewater but not in clinical cases is also captured in the logK analysis. It is apparent that this peak does not represent a new variant since the predominant variant at that point in time was Delta which emerged already in late spring 2021 (Fig. 3A). Possible explanations for this peak are explored in the Discussion section.

For Zurich, the arrival of Delta had a delayed effect on the increase in incidence, as the increase is observed slightly earlier in the viral load in wastewater as compared to reported cases (Fig. 3B). Interestingly, the emergence of BA.1 leads to an increase in incidence shown in a distinct peak based on reported cases, while this peak is much lower regarding the viral load in wastewater (Fig. 3B).

### 3.7 Estimation of variant abundance

We further aimed to estimate the abundance of VOCs by clustering SNPs over consecutive time windows of 28 days and taking the median of the SNP frequencies in each cluster for each time point (Fig. 6). For Altenrhein, the transition between Alpha and Delta is visible although it shows a strong noise level which can be explained with the poor quality of the wastewater data at this time period (Fig. 6A). This is in line with our previous findings for the Alpha/Delta transition from Swiss wastewater and with the poor estimation made by Freyja (Fig. 3). The take over of BA.1 over Delta is captured by our method, even though this transition takes places at the end of our time series for Altenrhein and there are fewer data points to average over leading to a noisier signal. For Geneva, the transitions from Delta to BA.1 and from BA.1 to BA.2 observed in clinical data are closely reproduced in our estimations which show distinctive signals (Fig. 6B). However, the transition signal from BA.2 to BA.5 is almost entirely missing. For Zurich, interestingly, we estimate Alpha to emerge earlier from wastewater than it is detected in clinical samples (Fig. 6C). Similar as for Altenrhein, the emergence of Delta in Zurich can be detected but is rather noisy. The transitions into BA.1, BA.2, and BA.5 have a clear, strong signal, reproducing the transitions obtained from clinical sampling very closely. To infer if there is any relation between the noise of the data and the accuracy of our abundance estimate we calculated a ranked correlation between the residuals of our estimate and the read counts, coverage and read depth, respectively. For all three catchments we see a significant negative correlation between the residuals of our estimate and the read coverage (Fig. S1, S2, S3) as determined by the Kendall’s Tau correlation coefficient (*R* = *−*0.22*, p <* 0.001 for Altenrhein, *R* = *−*0.22*, p <* 0.001 for Geneva, and *R* = *−*0.19*, p <* 0.001 for Zurich). A negative correlation was also observed between the read counts and the residuals for Altenrhein and Zurich but not for Geneva.

**Figure 6:**
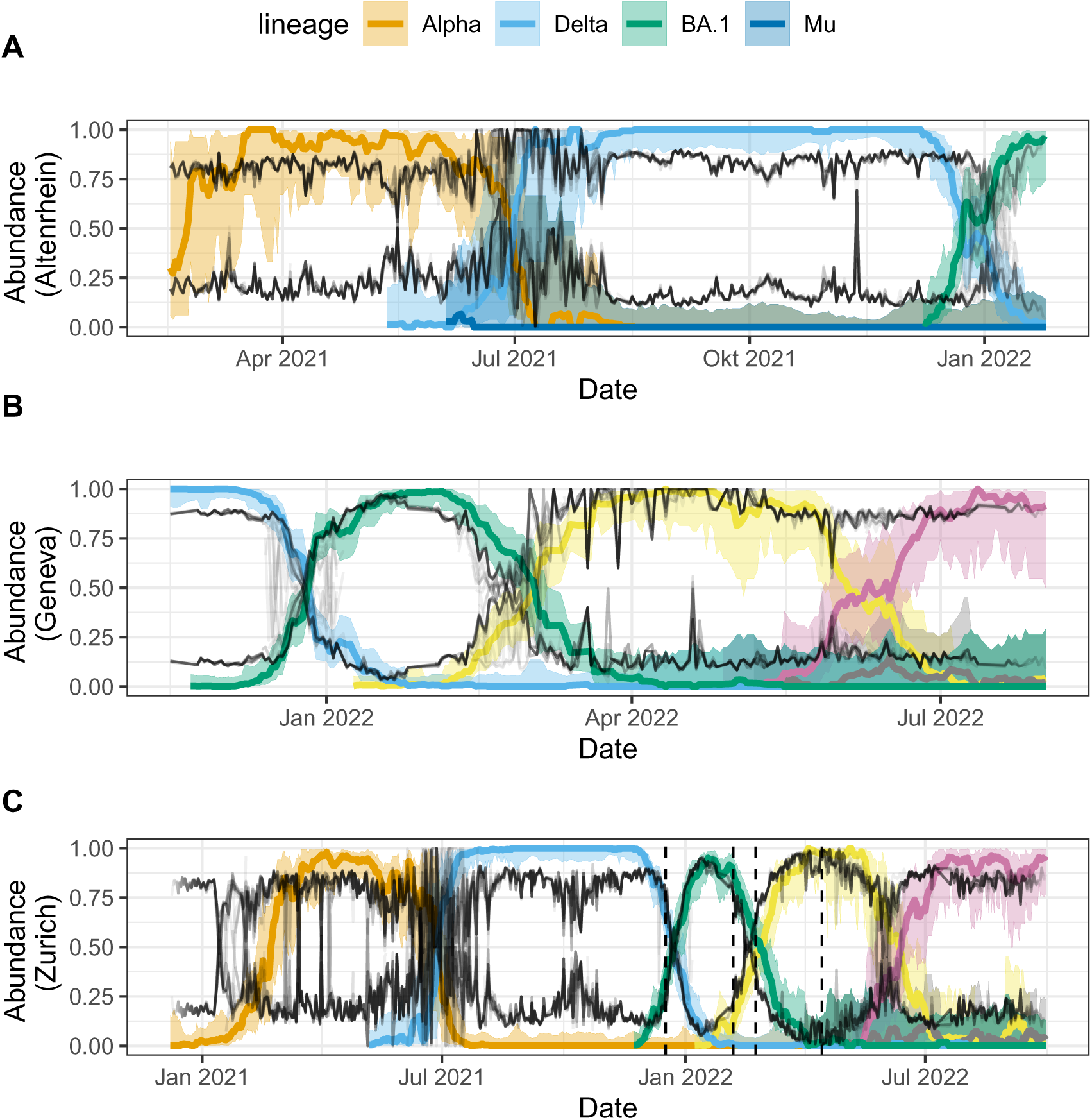
Abundance of VOCs for (A) Altenrhein, (B) Geneva, and (C) Zurich. The gray lines represent the estimated abundances when clustering SNPs over a time frame of 28 days, taking the median for each time point for each cluster and the mean of the medians for each time window.

## 4 Discussion

Here, we provide a novel approach using the genetic population structure as the basis for detecting new variants with increased fitness from wastewater samples taken on a nearly daily basis from three different Swiss wastewater catchment areas. We show that this method is able to distinguish the three main VOCs circulating at the time, Alpha, Delta, and Omicron, at their onset for all analyzed catchments independent of their population size or population density. The method was able to capture the emergence of a new lineage at an early stage in Geneva and Zurich, which are both catchments of large size, most likely due to a better signal to noise ratio. While a daily sampling strategy clearly gives the strongest signals, sampling three times a week is still sufficient to producing signals that correspond to the predominant variants.

Early detection of emerging variants using the proposed method critically depends on two main factors. Firstly the relative fitness gain of the variant over the contemporary mixture of lineages, and secondly the quality of the sampling and sequencing. Both affect the ratio between signal and noise, with the fitness gain increasing the signal and higher quality of sequencing data reducing noise. In the used dataset, this ratio is disadvantageous for the detection of both the Alpha and Delta variant, with relatively low read counts during the periods when, retrospectively, emergence of the variant is expected. For the Omicron variants, both fitness gain and data quality increase in all three locations, resulting in timely detection.

It should be pointed out that the close phylogenetic relatedness and close temporal succession of the Omicron BA sublineages [16] can lead to the method failing to produce a prominent distinction between those variants, as for instance visible in Geneva (Fig. SS5). Both temporal separation and earlier detection of the emerging variants can be achieved by using shorter time windows for the correlation analysis. However, shorter correlation analysis time windows by definition reduce the number of available data points, which reduces the chance of early detection when samples are not taken daily. Therefore, the correlation time should ideally be adapted to the sampling frequency.

Overall, a shorter correlation time might introduce more false positives but has the advantage of a higher sensitivity regarding sublineages and confined outbreaks. For example, we demonstrate that our method captures a peak in October 2021 in the Altenrhein wastewater, which coincided with an increase in viral load in the same samples, but was not observed as an increase in reported clinical samples. This deviation between wastewater and clinical samples is not surprising as the Altenrhein catchment represents a relatively small population and a few cases not caught by clinical testing can influence the wastewater incidence curve. An increase in incidence can stem from two different reasons: The takeover of a variant with a fitness higher than that of the predominant variant or a confined outbreak often caused by a superspreading event. As the two Delta sublineages AY.4 and AY.43 were the predominant lineage in the St.Gall area in October, we suspect that either of these two sublineages is responsible for a local COVID-19 outbreak in the Altenrhein catchment area. As both sublineages were prevalent in the whole of Europe and beyond in autumn 2021, we reason that both lineages do not have an increased fitness advantage over other circulating Delta sublineages. Interestingly, lineage AY.43 also reemerged during an Omicron wave in two wastewater treatment plants in the Netherlands during the months of August and September of 2022 [15].

When a new, more fit lineage emerges and logK reaches a certain threshold the fitness advantage of the predominant set of SNPs stemming from this lineage can be used to estimate the current abundance of the new lineage using k-means clustering. While we show that the k-means clustering of SNPs gives a good estimate of the abundance overall, when sequencing data is of insufficient quality the estimate becomes noisy. In particular, we observe an inverse correlation between the residuals of our abundance estimate and the read counts. However, when samples are taken on a regular basis and sufficient sequencing quality is given, the abundance can be estimated to a degree where the growth rate of the new variant can be estimated. We believe that this approach is particular useful for areas that lack access to sufficient resources for sequencing of clinical samples as the whole pipeline only relies on wastewater sampling and sequencing. These methods can complement already established practices that use wastewater to estimate incidence and *R*_0_.

Other methods for detecting lineages in mixed samples, such as Freyja, are able to detect the emergence of new variants earlier than our method. However, it must be noted that those estimates have been conducted retrospectively after the predominant lineages of this time period have already been defined, as Freyja relies on a pre-definition of lineages for its valuation, an estimation in real time could therefore experience some delay. The here proposed method is agnostic of the defined lineages, and therefore is able to detect completely unknown lineages with increased fitness without human intervention. Previously one study analyzed the genetic population structure based on Yue and Clayton measure of dissimilarity index at each position using it as a measure for the distance between samples [11]. While both approaches share the main idea, the strength of our approach lies in its time resolved prediction of upcoming variants. The fact that our method is based on time resolution also makes it robust against outliers.

One drawback when using wastewater for surveillance could be that different variants have different shedding rates which might influence the viral load and the amount of genomic material found in wastewater. One advantage of using logK is that it considers the relative change in genetic diversity and is therefore proportionally related to the growth rate of the new lineage, while the growth rate itself is not dependent on the shedding rate of the specific variant [17]. Thus, shedding is not expected to bias the variant estimation using our metric. However, shedding can have an effect on the abundance and *R*. This effect is larger with increased growth rate, therefore the effect might be higher for the Omicron variant than for Delta [17].

One must also consider that the detection of lineages from wastewater can be distorted by long-term infections of immunocompromised patients as well as animal reservoirs [23]. These factors can also influence the viral load gained from wastewater leading to differences in incidence between estimations based on wastewater and reported cases, as it is the case for Zurich during the emergence of BA.1. However, our method using the genetic diversity is robust against those fluctuations as it only relies on changes in the genetic population structure between two time points and not on the absolute viral load.

We recognize that there are several parameters that influence the course of logK. For example the height of the logK peak is proportional to the genetic diversity between the predominant and the emerging lineage, i.e. the difference in the number of mutations between the variants. As the genetic distance between Delta and Omicron is larger than between Alpha and Delta, the peak signalling Omicron is higher in value. Consequently, lineages that are particularly distant, for example due to recombination, are easier to detect. Furthermore, we rely on the pre-definition of a time period for correlation calculation to estimate p-values and a detection threshold which must distinguish the signal from noise. Furthermore, as logK is calculated between the current and a previous date and the difference in logK is averaged over a past time period to capture its trend, some amount of historical data is required. This also explains the difficulty of estimating a variant in cases where it occurred in the beginning of the analyzed time periods, as is the case for Alpha in Altenrhein.

We conclude that the logK-based variant detection method is able to reliably detect emerging viral variants in wastewater samples, if the quality criteria are met. Samples need to be taken at least every other day, preferably every day while adhering to a strict sampling protocol, and sequenced to sufficient depth. Under these conditions, this method can be used as part of wastewater surveillance systems when monitoring for new lineages with a higher fitness than the circulating lineage. Because the proposed method does not require prior lineage classification, as opposed to the most commonly used methods, it is able to detect every emerging variant, independent of their identification in clinical isolates.

## Data Availability

All data produced in the present study are available upon reasonable request to the authors

