## Supplemental Material for "Wastewater surveillance without prior lineage classification for reliable real time SARS-CoV-2 variant identification"

### Supplementary Information

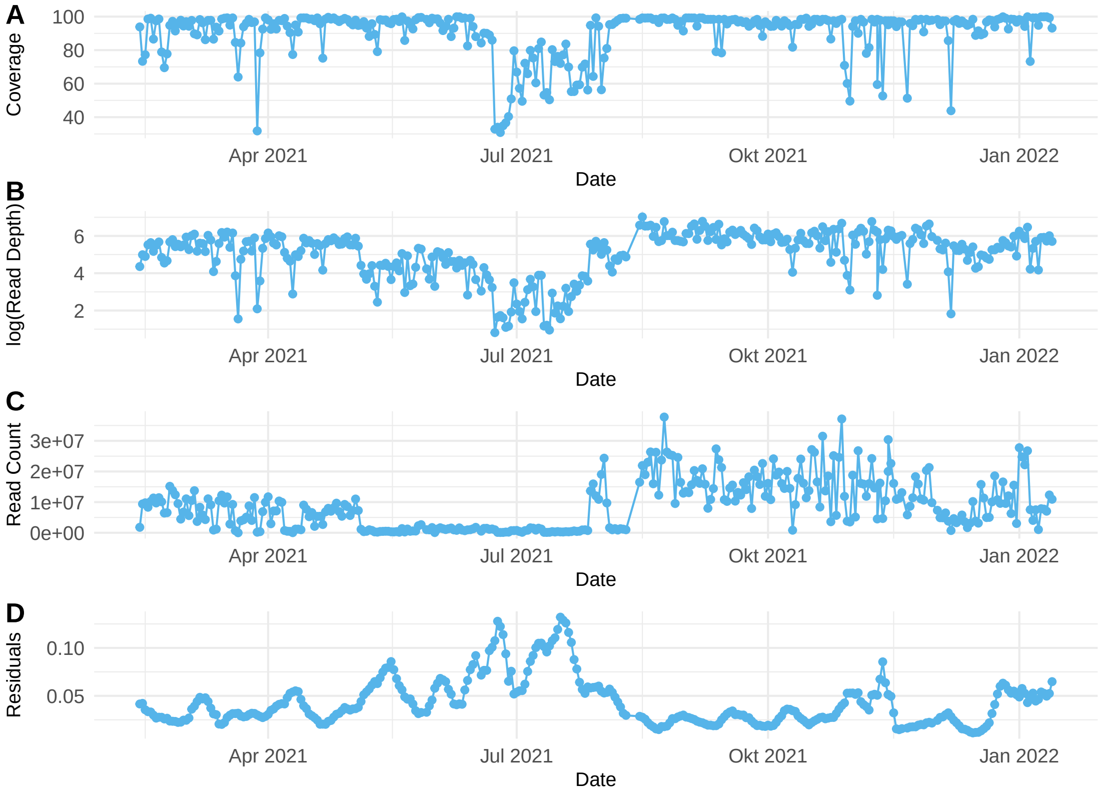


Figure S1: Quality assessment of sequencing data for Altenrhein. The coverage (A), log(read depth) (B), and read counts (C) over the analyzed time period are depicted. (D) shows the residuals from the abundance estimation based on a rolling average over 25 days.

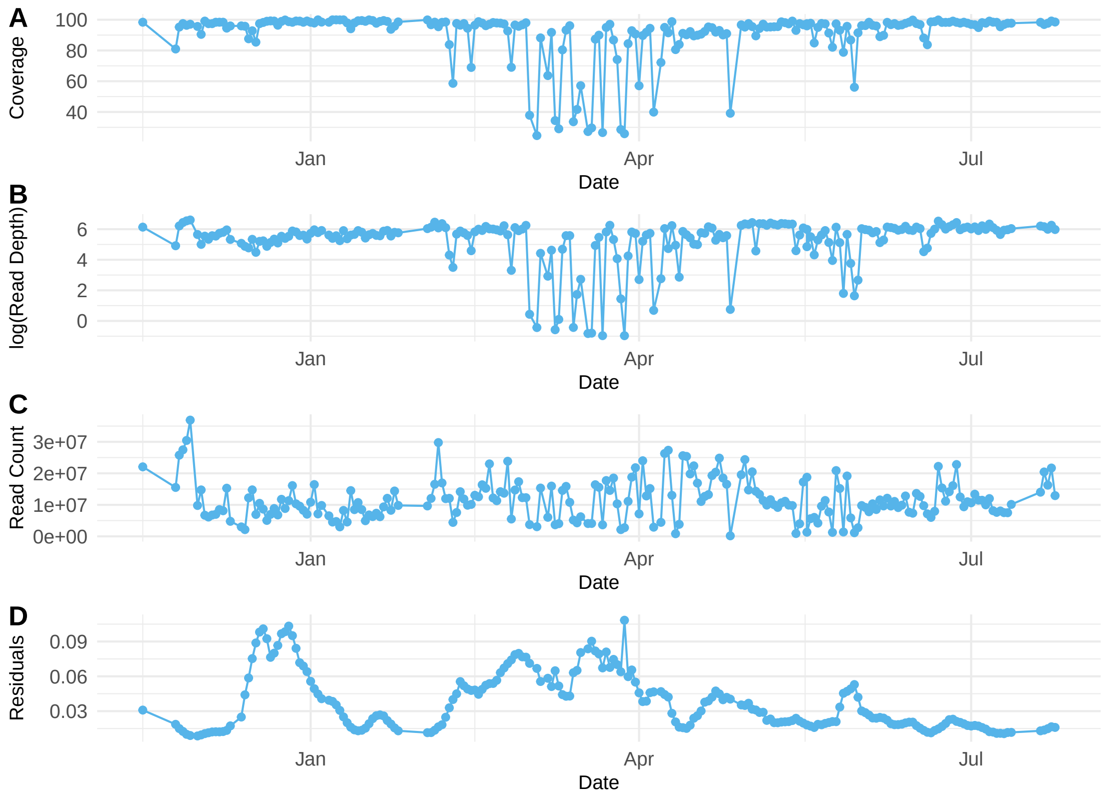


Figure S2: Quality assessment of sequencing data for Geneva. The coverage (A), log(read depth) (B), and read counts (C) over the analyzed time period are depicted. (D) shows the residuals from the abundance estimation based on a rolling average over 25 days.

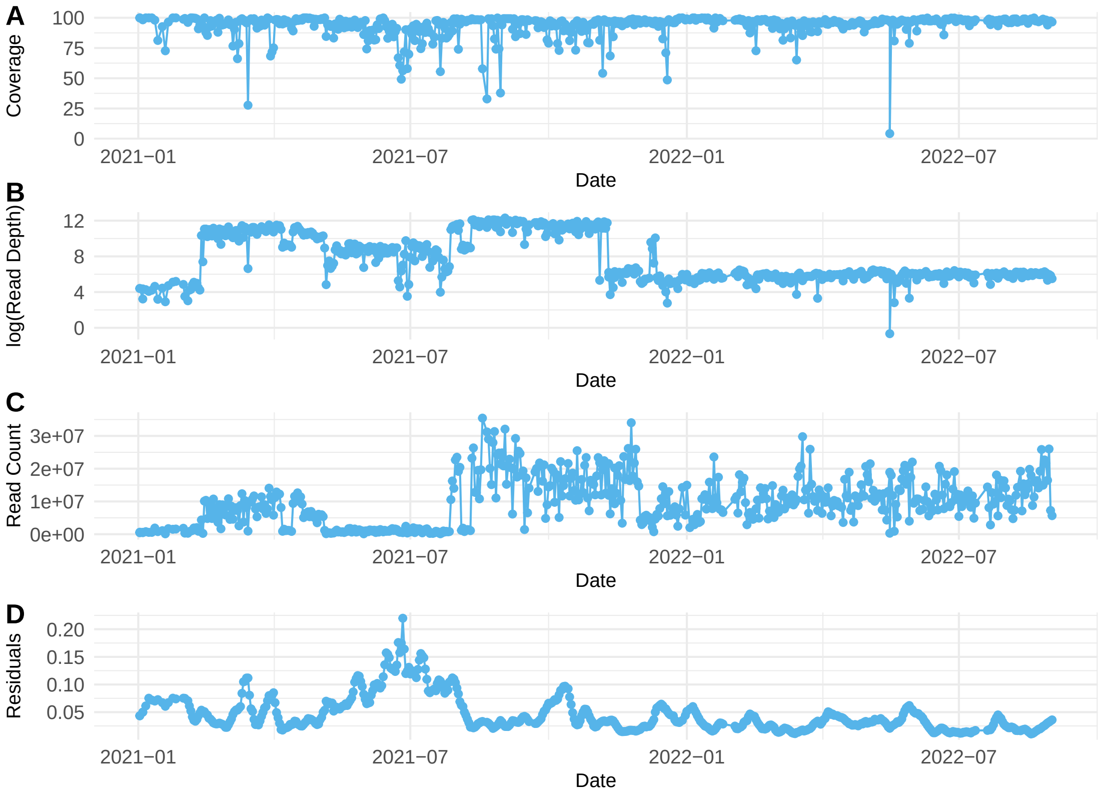


Figure S3: Quality assessment of sequencing data for Zurich. The coverage (A), log(read depth) (B), and read counts (C) over the analyzed time period are depicted. (D) shows the residuals from the abundance estimation based on a rolling average over 25 days.

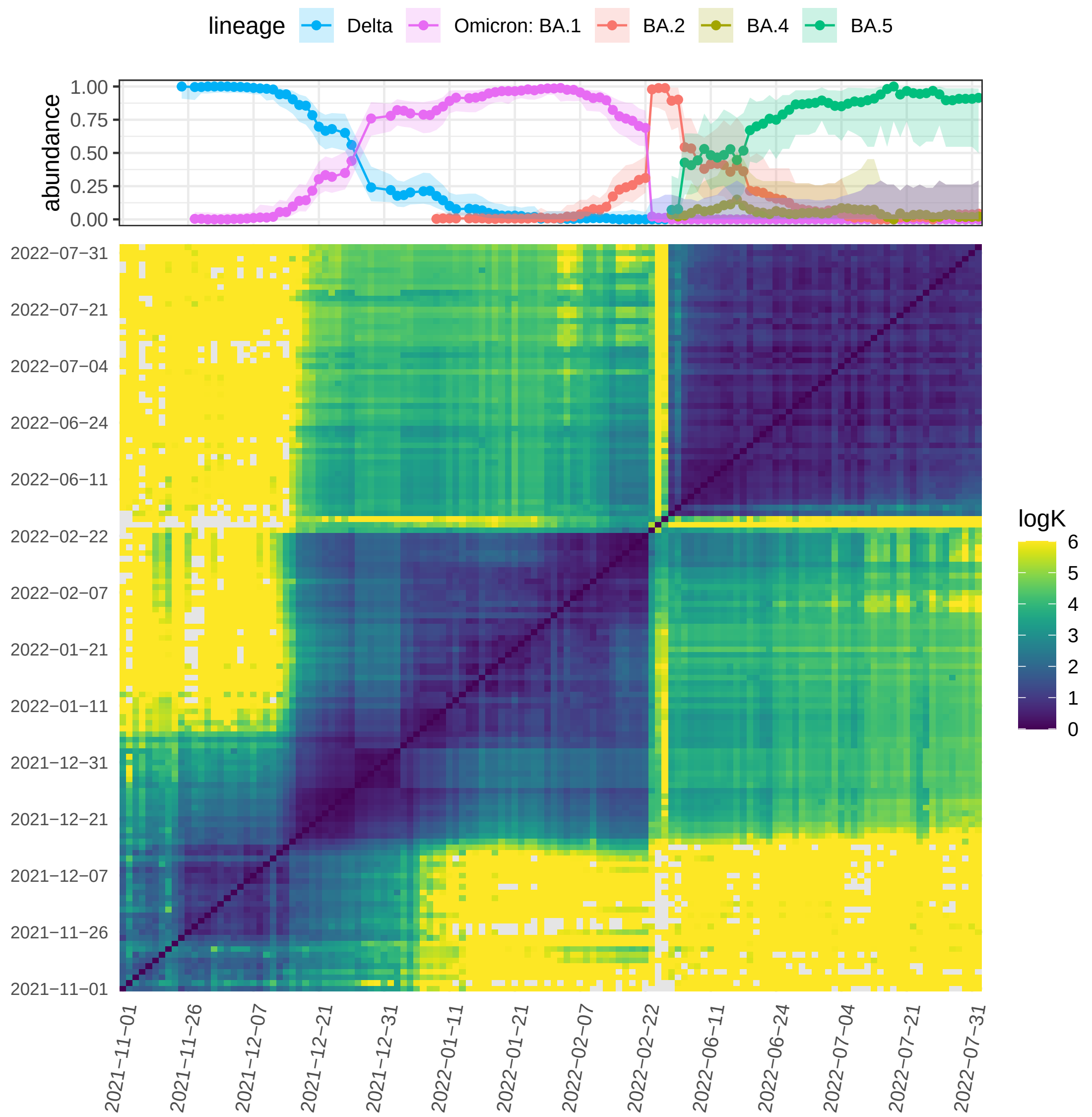


Figure S4: logK estimates on an nearly daily basis for Geneva for all date pairs. Above the abundance of VOCs based on clinical cases taken from GISAID is shown. Abundance estimates are shown only for days on which wastewater samples are available and displayed on a discrete time scale.

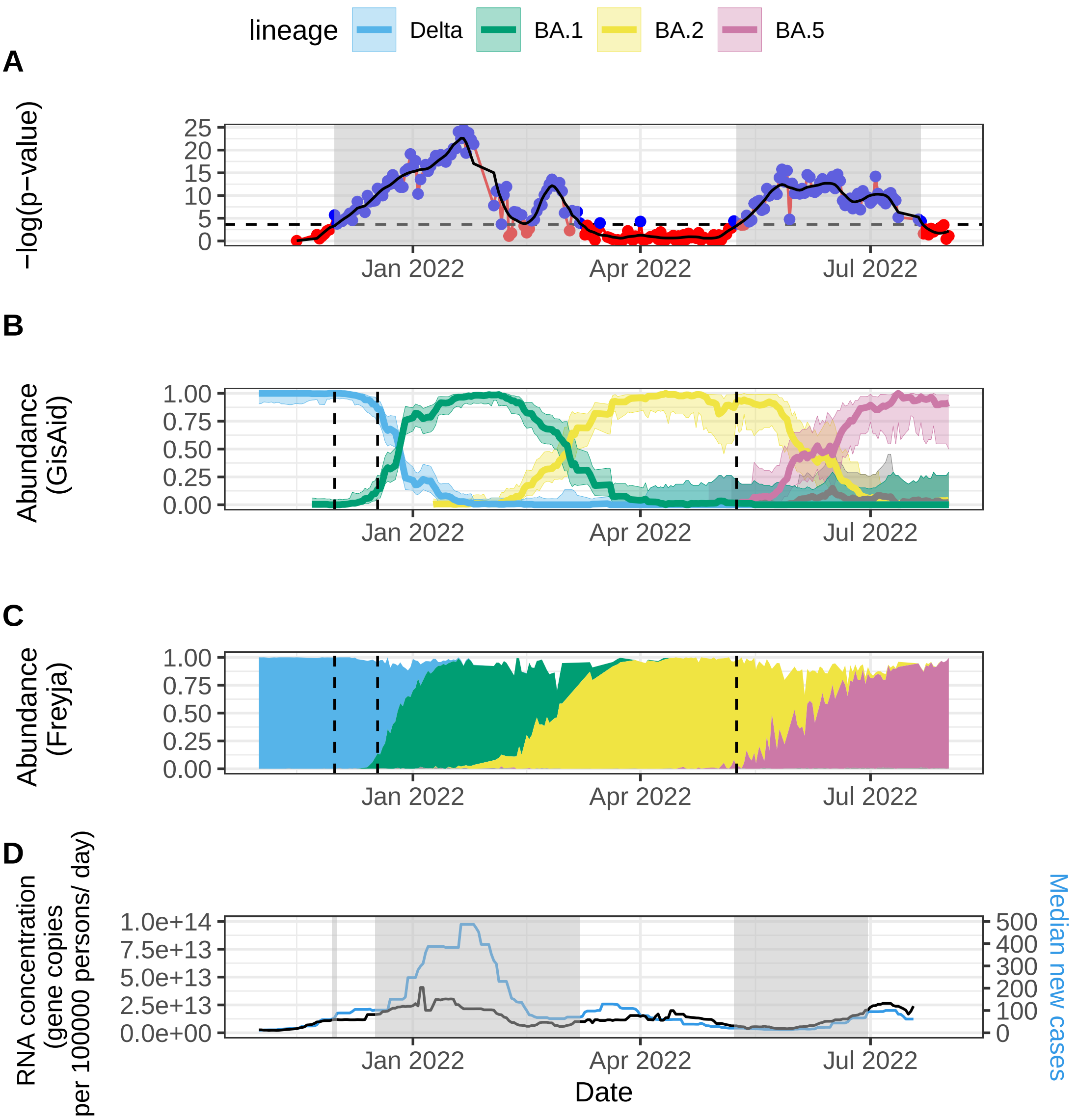


Figure S5: Variant and incidence estimates for Geneva. (A) P-values for logK ranked correlations with time for periods of 42 days. The black line represents loess smoothing. Blue dots and grey transparent areas represent values that are above the detection threshold (see Methods section, dashed horizontal line). (B) Abundance of VOCs based on clinical samples obtained from GISAID. (C) Estimation of VOCs in local wastewater using Freyja. (D) SARS-CoV-2 RNA concentration in the local wastewater facility as a function of time and the median local case incidence.

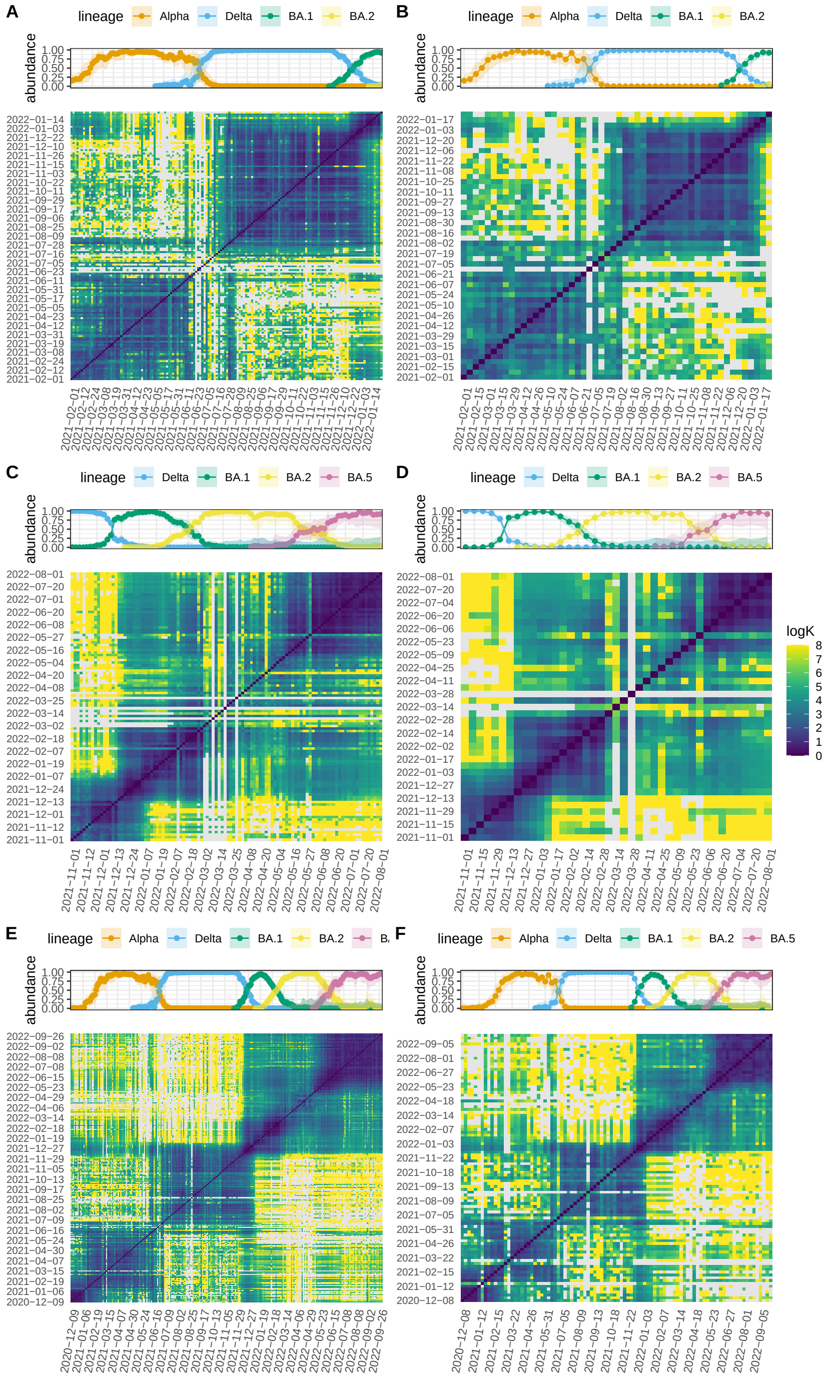


Figure S6: logK estimates based on sequence data from wastewater for Altenrhein, St Gall (A, B), Geneva (C, D), and Zurich (E, F) for all date combinations. Left panel (A, C, E) represents logK values for samples selected three times a week while right panel (B, D, F) depicts logK values for weekly samples. Above each plot the abundance of VOCs based on clinical cases taken from GISAID is shown. Only dates on which wastewater samples are available were selected and displayed on a discrete timeline.

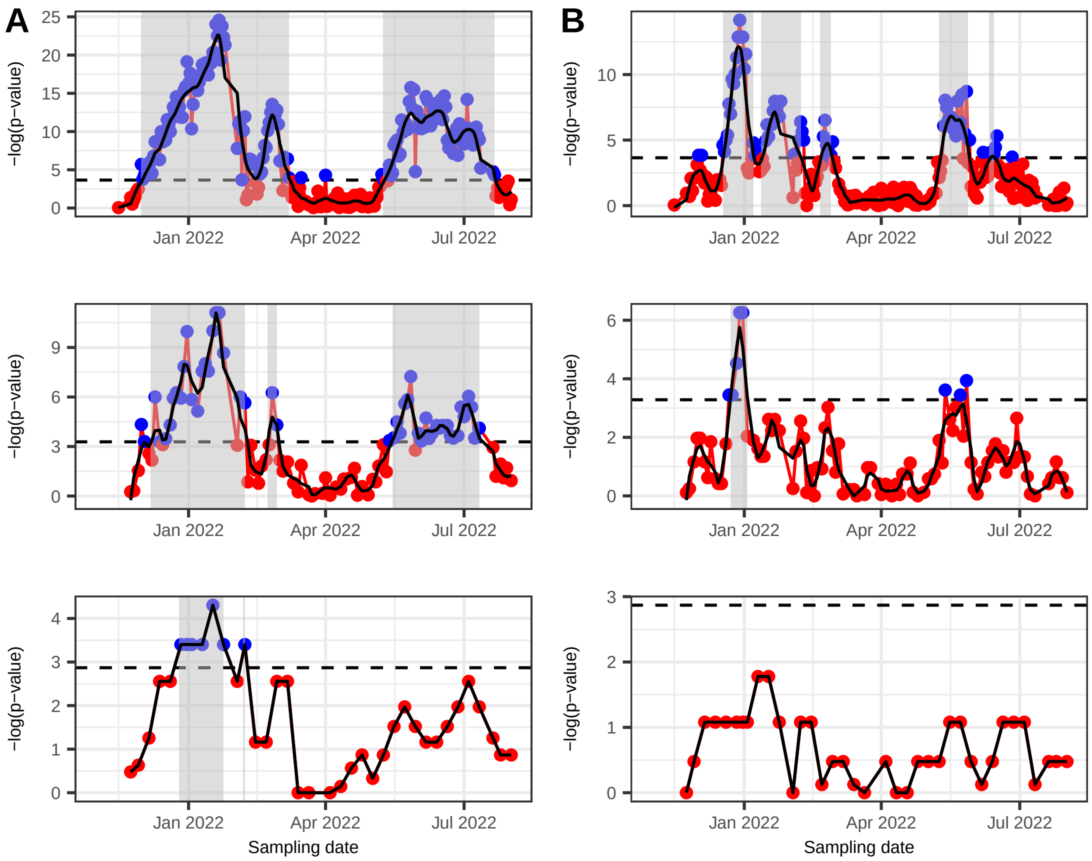


Figure S7: Sensitivity analysis of p-values for the logK correlations for Geneva. The correlations were performed over 42 (A) and 21 (B) days. The panels represent correlations for samples taken daily (upper panel), three times a week (middle panel), and weekly (lower panel). The dashed horizontal line represents the threshold for detection (see Methods section).

Table 1: **First detections of VOCs in Altenrhein wastewater. Columns "first date" and "last date" represent the start and end day of the predominant variant or lineage defined based on clinical data (See method section for details). The remaining columns show the days of detection when sampling is carried out daily, three times a week, or weekly. Estimated detection dates are based on p-values for logK correlations of 21 days.**

| lineage | first_date | last_date | daily_offset21 | MoWedFri_offset21 | weekly_offset21 | daily_point_esti | MoWedFri_point_esti | weekly_point_esti |
|---|---|---|---|---|---|---|---|---|
| Alpha | 2021-02-16 | 2021-03-17 | 2021-04-24 | NA | NA | 2021-02-18 | NA | NA |
| Delta | 2021-06-10 | 2021-07-09 | 2021-10-21 | NA | NA | 2021-06-16 | NA | NA |
| Omicron: BA.1 | 2021-12-11 | 2022-01-18 | 2021-12-25 | 2022-01-03 | NA | 2021-12-23 | 2022-01-03 | 2022-01-17 |

Table 2: **First detections of VOCs in Zurich wastewater. Columns "first date" and "last date" represent the start and end day of the predominant variant or lineage defined based on clinical data (See method section for details). The remaining columns show the days of detection when sampling is carried out daily, three times a week, or weekly. Estimated detection dates are based on p-values for logK correlations of 21 days.**

| lineage | first_date | last_date | daily_offset21 | MoWedFri_offset21 | weekly_offset21 | daily_point_esti | MoWedFri_point_esti | weekly_point_esti |
|---|---|---|---|---|---|---|---|---|
| Alpha | 2021-01-23 | 2021-03-30 | NA | NA | NA | NA | NA | NA |
| Delta | 2021-06-08 | 2021-07-14 | NA | NA | NA | NA | NA | NA |
| Omicron: BA.1 | 2021-12-02 | 2022-01-24 | 2021-12-24 | 2021-12-27 | NA | 2021-12-19 | 2021-12-17 | NA |
| BA.2 | 2022-01-24 | 2022-04-02 | 2022-03-03 | NA | NA | 2022-02-15 | NA | NA |
| BA.5 | 2022-05-17 | 2022-07-19 | NA | NA | NA | 2022-05-24 | 2022-05-30 | NA |

Table 3: **First detections of VOCs in Geneva wastewater. Columns "first date" and "last date" represent the start and end day of the predominant variant or lineage defined based on clinical data (See method section for details). The remaining columns show the days of detection when sampling is carried out daily, three times a week, or weekly. Estimated detection dates are based on p-values for logK correlations of 42 days.**

| lineage | first_date | last_date | daily_offset42 | MoWedFri_offset42 | weekly_offset42 | daily_point_esti | MoWedFri_point_esti | weekly_point_esti |
|---|---|---|---|---|---|---|---|---|
| Delta | 2021-11-16 | 2021-11-16 | NA | NA | NA | NA | NA | NA |
| Omicron: BA.1 | 2021-12-13 | 2022-01-18 | 2021-12-02 | 2021-12-08 | 2021-12-27 | 2021-12-13 | 2021-12-27 | NA |
| BA.2 | 2022-02-09 | 2022-03-28 | 2022-02-15 | 2022-02-23 | 2022-02-07 | 2022-02-10 | NA | NA |
| BA.5 | 2022-05-16 | 2022-07-11 | 2022-05-10 | 2022-05-16 | NA | 2022-05-16 | NA | NA |

Table 4: **First detections of VOCs in Geneva wastewater. Columns "first date" and "last date" represent the start and end day of the predominant variant or lineage defined based on clinical data (See method section for details). The remaining columns show the days of detection when sampling is carried out daily, three times a week, or weekly. Estimated detection dates are based on p-values for logK correlations of 21 days.**

| lineage | first_date | last_date | daily_offset21 | MoWedFri_offset21 | weekly_offset21 | daily_point_esti | MoWedFri_point_esti | weekly_point_esti |
|---|---|---|---|---|---|---|---|---|
| BA.1 sublineage (?) | NA | NA | 2021-12-19 | NA | NA | NA | NA | NA |
| BA.2 | 2022-02-09 | 2022-03-28 | 2022-02-21 | NA | NA | 2022-02-09 | NA | NA |
| BA.5 | 2022-05-16 | 2022-07-11 | 2022-05-10 | NA | NA | 2022-05-16 | 2022-05-23 | NA |
| BA.5 sublineage (?) | NA | NA | 2022-06-12 | NA | NA | NA | NA | NA |
